# The association between docosanoic acid and the risks of occurrence and mortality of chronic kidney disease

**DOI:** 10.1101/2025.03.14.25322644

**Authors:** Xianglong Meng, Qianyu Yang, Zhuoxing Li, Ping Zhou, Wan Li, Qiyuan Liang, Tingyu Wu, Wuyu Gao, Haiyan Yu, Guifei Deng, Junlin Zhang, Xiang Xiao

## Abstract

**Objective:** This study aims to investigate the association between docosanoic acid (DA) in human circulation and the risk of occurrence and mortality of chronic kidney disease (CKD).

**Methods:** This is a cross-sectional study including individuals in NHANES 2011-2014. RCS were used to fit the dose-effect curve between DA levels and CKD risk in the general population, and mortality in CKD patients. Multiple logistic and Cox regressions were used to analyze the association of DA with CKD occurrence and mortality risks respectively.

**Results:** A total of 2,366 participants were included in this study, including 1,958 (82.8%) individuals without CKD and 408 (17.2%) CKD individuals. The RCS results showed a linear association between DA and the risk of CKD in individuals. Per standard deviation (per-SD) increase in DA, the risk of CKD in the general population decreased by 18% (OR = 0.82; 95% CI, 0.70 - 0.96; *P* = 0.02). In addition, the RCS results showed a linear association between DA and the risk of cardiovascular and all-cause mortality in CKD. Multivariate Cox regression analysis showed that per-SD increase in DA, the risk of cardiovascular mortality in CKD patients decreased by 44% (HR = 0.56; 95% CI, 0.35 - 0.90; *P* = 0.02) and all-cause mortality decreased by 27% (HR = 0.73; 95% CI, 0.59 - 0.89; *P* = 0.002).

**Conclusion:** Higher serum DA in populations means lower CKD risk. Moreover, for CKD patients, as DA levels increase, the risk of cardiovascular and all-cause mortality decreases.

## 1. Introduction

Chronic kidney disease (CKD) is a debilitating disease that affects 10 - 15% of the adult population worldwide. From 2022 to 2027, the global prevalence of CKD will continue to increase. It is estimated that by 2027, the total prevalence of CKD will rise to 436.6 million (an increase of 5.8% compared to 2022) ^1^. If there is no active intervention, it may cause a huge clinical burden ^1^.

Docosanoic acid (DA) is a straight-chain, C22 long-chain saturated fatty acid and belongs to very long-chain fatty acids (VLCFA) ^2^. DA naturally exists in many essential oils, among which peanuts, macadamia nuts and rapeseed oil have the highest total amount of VLCFA ^3^. Among the major commercial oils, sunflower oil has a higher content of 22:0 (DA) ^4^. Fatty acids, as components of animal and plant fats, are widely distributed in nature and are an important part of the normal daily diet of mammals, birds and invertebrates. Existing studies have shown that DA may play a certain role in some cardiovascular diseases and also has potential relations with the pathological processes of certain metabolic diseases. The study by Fretts AM et al. ^5^ indicated that the higher the circulating concentration of DA, the lower the risk of diabetes. In addition, higher circulating DA levels are associated with a lower risk of atrial fibrillation (AF) ^6^. At the same time, studies have also shown that high-level circulating VLCFA (C22:0 and C24:0) have protective effects on cardiovascular diseases, coronary heart disease and all-cause mortality in the entire population, the hyperlipidemia population and the hypertensive population ^7^.

Moreover, DA may be involved in certain anti-inflammatory and antioxidant mechanisms ^8,9^. Meanwhile, inflammation and oxidative stress have been confirmed to play important roles in the occurrence and development of CKD ^10,11^. However, the specific association between it and CKD is still unclear.

Therefore, this study aims to explore the association between DA and the risk of occurrence and mortality of CKD. This may provide new biomarkers for screening people at high risk of CKD and also for patients with CKD who are at high risk of incidence of mortality.

## 2. Materials and Methods

### 2.1 Study Design and Individuals

The National Health and Nutrition Examination Survey (NHANES) is a continuous study which gathers nutrition and health-related data for adults and children in the United States ^12^. NHANES intends to probe into individual-level demographic, health and nutrition information by means of personal interviews and standardized physical examinations carried out at specialized Mobile Examination Centers (MECs), and to evaluate the health and nutritional status of non-institutionalized civilians in the United States ^13^. The relevant mortality data were retrieved from the National Mortality Index (NDI) database of the Centers for Disease Control and Prevention (CDC) in the United States. The data utilized in this study were sourced from the National Nutrition and Health Examination Survey (NHANES) database spanning from 2011 to 2014.

To ensure the accuracy of the study, specific criteria were used to define or diagnose the presence of smoking, alcohol use, diabetes, hypertension, hyperlipidemia, and anemia **(Supplementary - table 1)**. Circulating DA levels were detected by electron capture negative ion mass spectrometry. Individuals with an albumin-to-creatinine ratio (ACR) higher than 30 mg/g and/or an estimated glomerular filtration rate (eGFR) lower than 60 mL/min/1.73 m^2^ were defined as CKD patients ^14^.

The exclusion criteria for this study were: 1) age less than 20 years, 2) pregnancy, 3) loss of CKD diagnosis, 4) dialysis, and 5) missing DA data **(Figure 1)**. The study protocol conformed to the ethical standards of the Declaration of Helsinki of 1964 and its subsequent amendments. All procedures involving human participants were approved by the Research Ethics Review Board of the National Center for Health Statistics Research, and all participants signed informed consent forms ^15^.

**Figure 1.**
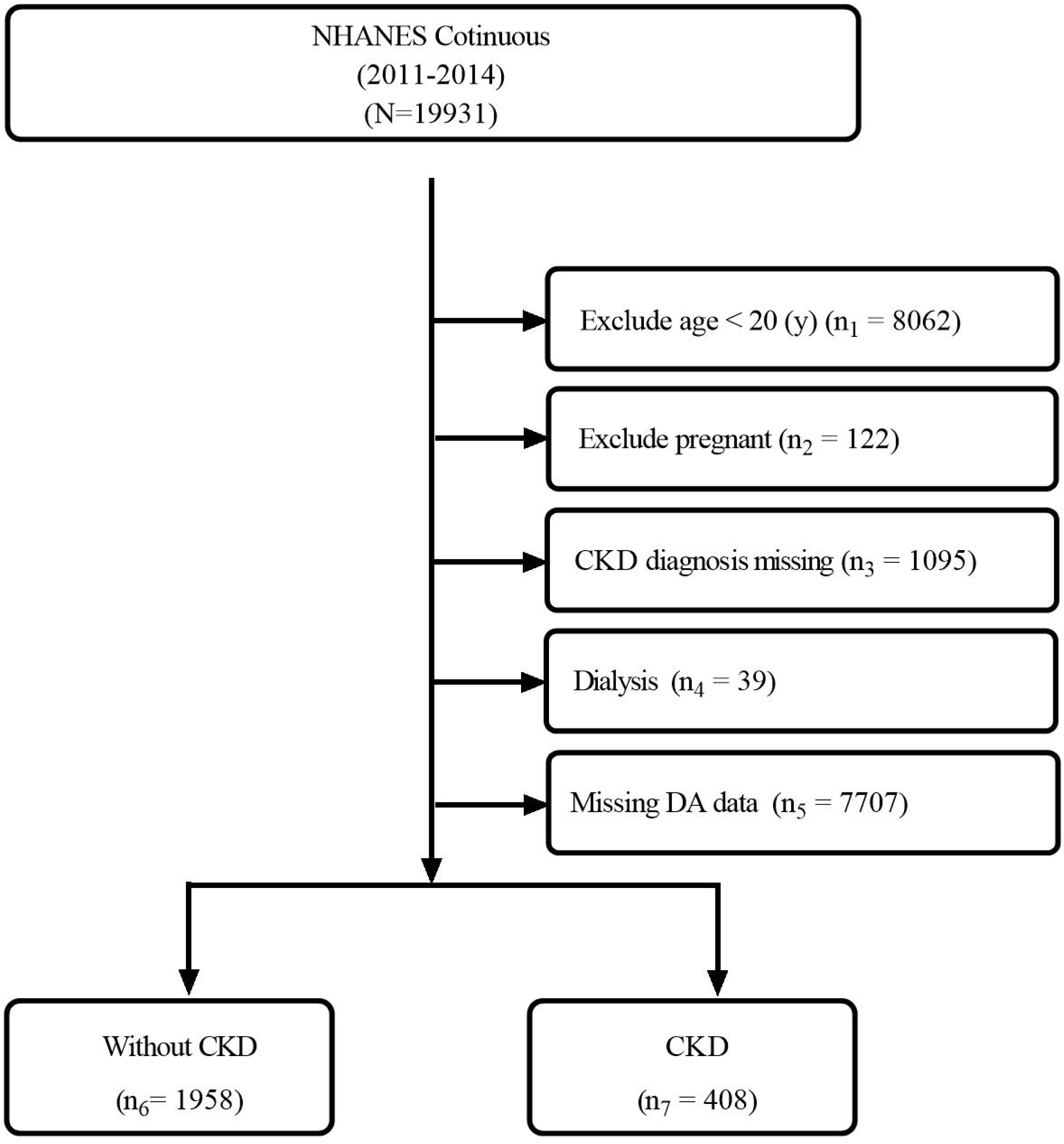

### 2.2 Statistical Analysis

According to the CDC guidelines, all computational analyses took weighted samples into account and designed stratification and clustering to obtain estimates for the general population of the United States. For categorical variables, they were described by weighted percentages (standard errors), and for continuous variables, baseline characteristics were described by weighted means (standard errors). Complex sampling restricted cubic spline (RCS) plots were used to assess the association between DA levels and the risk of CKD occurrence and mortality. A weighted multivariate Logistic regression model was used to analyze the association between DA and the risk of CKD occurrence in the general population, and the consistency of the results was evaluated through stratified analysis. In the sensitivity analysis, multiple imputation was performed on the missing data and DA was grouped by the trisection method, and then multivariate Logistic regression analysis was conducted to observe the consistency of the results. A multivariate Cox regression model was used to analyze the association between DA levels and the risk of mortality in CKD patients. The calculation method of the weights was to determine the smallest subset based on the variables included in the study and select the corresponding weights. Finally, the weights over the years were combined. All statistical tests were performed using R 4.2.2, and a two-sided P < 0.05 was considered statistically significant.

## 3. Results

### 3.1 Baseline Characteristics

In this study, we included 19,931 individuals enrolled in the NHANES database from 2011 to 2014. After exclusion based on the exclusion criteria, a total of 2,366 participants were finally included, among which 1,958 (82.8%) individuals did not have CKD and 408 (17.2%) individuals had CKD **(Figure 1)**. After weighting, the average age of all individuals was 48.05 years, 49.01% were male, and the average DA level was 68.56 umol/L **(Table 1)**.

### 3.2 Association between DA level and the risk of individual CKD occurrence

#### 3.2.1 The risk of CKD occurrence is lower in patients with higher DA levels

The individual was divided into a low DA level group (≤ 65.10 umol/L) (n_1_ = 1,185) and a high DA level group (> 65.10 umol/L) (n_2_ = 1,181) using the dichotomy method **(Table 1)**. Compared with the lower DA group, individuals in the higher DA group were older, had more females, had a relatively higher education, had more non-Hispanic blacks and non-Hispanic whites, and had fewer Mexican Americans and other Hispanics (All *P* < 0.05). Individuals in the higher DA group had a higher serum albumin, a lower proportion of anemia and diabetes; individuals in the higher DA group had a relatively larger BMI, and a higher proportion of individuals with hypertension and hyperlipidemia (All *P* < 0.05). In addition, in the baseline data, individuals in the higher DA level group had a higher glomerular filtration rate (eGFR), a lower urinary albumin creatinine ratio (ACR), and less use of renin-angiotensin-aldosterone system inhibitors (RAASi) than individuals in the lower DA group (All *P* < 0.05) **(Table 1)**.

RCS was used to fit the dose-effect association between DA and the risk of CKD occurrence in the general population. The results showed that the individual DA level was linearly related to the risk of CKD occurrence. As the DA level increased, the risk of individual CKD occurrence decreased (nknot = 5, Non-line *P* = 0.63) **(Figure 2)**. After adjusting for baseline variables such as age, sex, ethnicity, BMI, smoking (“yes” or “no”), alcohol use (“yes” or “no”), poverty, education, anemia (“yes” or “no”), hyperlipidemia (“yes” or “no”), hypertension (“yes” or “no”), diabetes (“yes” or “no”), RAASi use (“yes” or “no”), the results of multivariate Logistic regression analysis showed that compared with individuals with lower DA, individuals with higher DA levels had a 42% lower risk of CKD (OR: 0.58; 95% CI, 0.39 - 0.87; *P* = 0.01). In addition, per standard deviation (per-SD) increment of circulating DA was associated with an 18% reduction in the risk of CKD occurrence (OR = 0.82; 95% CI, 0.70 - 0.96; *P* = 0.02). **(Figure 3, Supplementary - table 2 and 3)**

**Figure 2.**
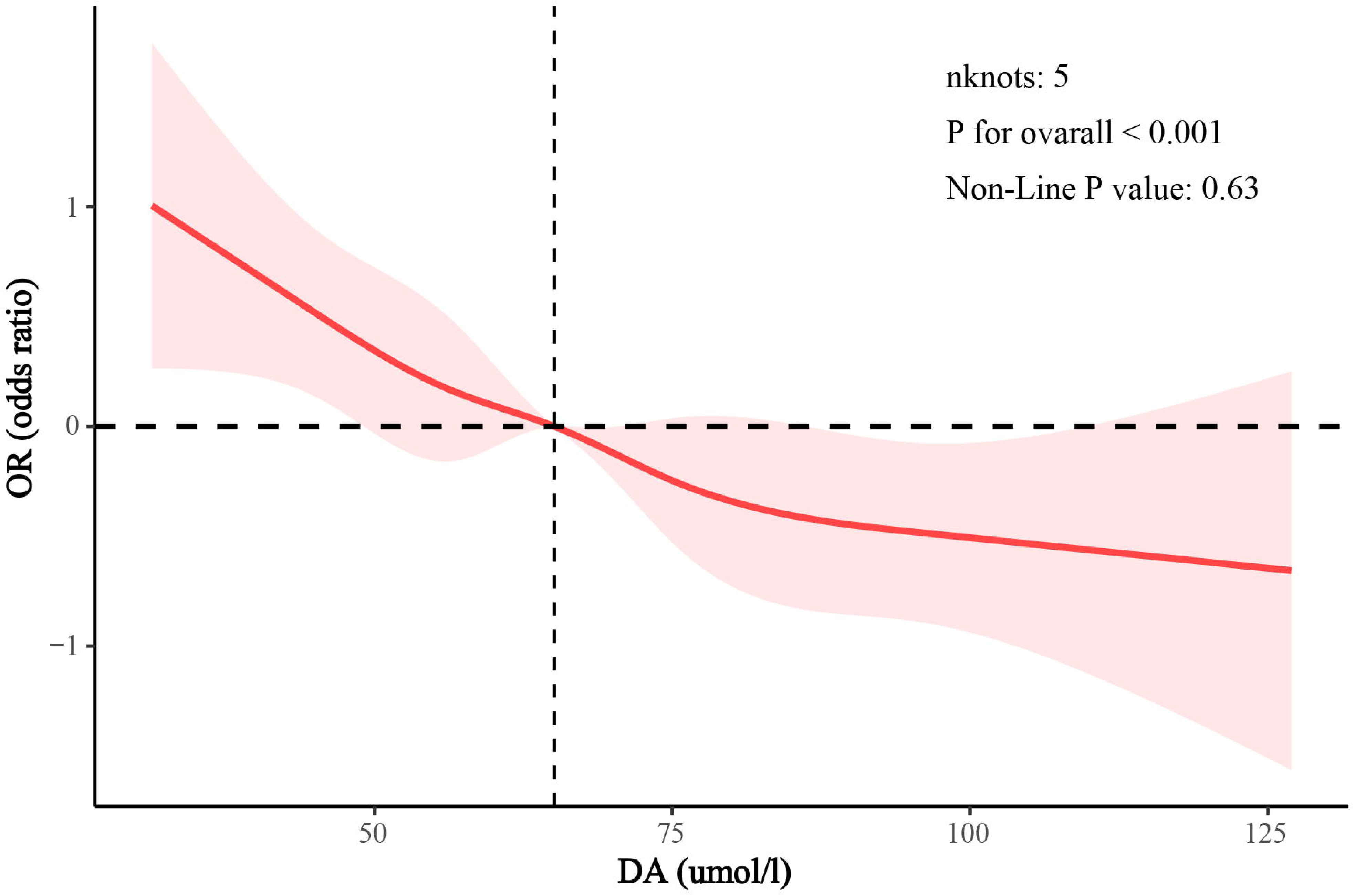

**Figure 3.**
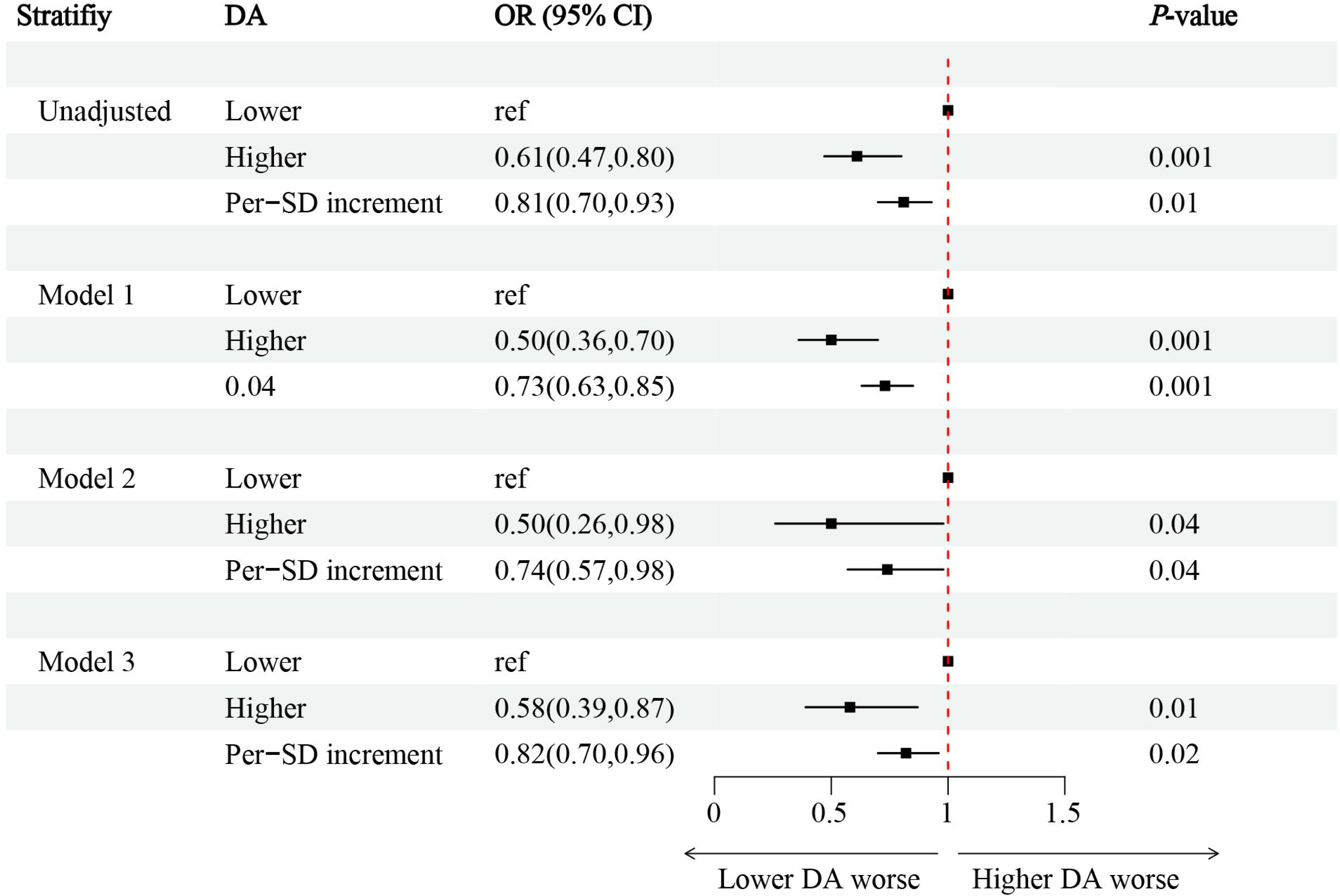

#### 3.2.2 Sensitivity analysis

Based on the differences in individual age, sex, ethnicity, BMI, anemia (“yes” or “no”), hypertension (“yes” or “no”), diabetes (“yes” or “no”), etc., we conducted stratified analysis. The results showed that at different levels, this correlation between higher DA and lower risk of CKD occurrence was consistent (All *P* for interaction > 0.05) **(Supplementary - table 4)**.

To eliminate the influence of missing values on the research results, we performed multiple imputations on the data. After data imputation, the results of multivariate Logistic regression analysis showed that compared with individuals with lower DA, individuals with higher DA had a 39% lower risk of CKD occurrence (OR = 0.61; 95% CI, 0.43 - 0.88; *P* = 0.01) (Supplementary - table 5). In addition, we grouped individuals with different DA levels using the tertile method. The results of multivariate Logistic regression analysis showed that compared with individuals in the T1 group, individuals in the T3 group had a 36% lower risk of CKD (OR = 0.64; 95% CI, 0.43 - 0.95; *P* = 0.03) **(Supplementary - table 6)**.

### 3.3 Association between DA level and the risk of mortality in CKD patients

Among the 408 included CKD patients, the average age after weighting was 60.52 years, of which 43.16% were male, the average eGFR was 75.38 ml/min/1.73^2^, and the average ACR was 167.65 mg/g. The average level of weighted DA was 65.66 umol/L, and the proportion of mortality was 30.88% **(Supplementary - table 7)**.

The RCS fit the association between DA and the risk of cardiovascular and all-cause mortality in CKD patients. The results showed that there was a linear association between DA level and the risk of cardiovascular mortality in CKD patients (nknot = 5, Non-line *P* = 0.57). As the DA level increased, the risk of cardiovascular mortality in CKD patients increased **(Figure 5. A)**. There was also a linear relationship between DA level and the risk of all-cause mortality in CKD patients (nknot = 5, Non-line *P* = 0.94). As the DA level increased, the risk of all-cause mortality in CKD patients also increased **(Figure 5. B)**. The results of multivariate Cox regression analysis showed that for per-SD increase in DA level, the risk of cardiovascular mortality in CKD patients decreased by 44% (HR = 0.56; 95% CI, 035 - 0.90; *P* = 0.02) and the risk of all-cause mortality decreased by 27% (HR = 0.73; 95% CI, 0.59 - 0.89; *P* = 0.002) **(Table 2, Supplementary - table 8, 9)**.

**Figure 4.**
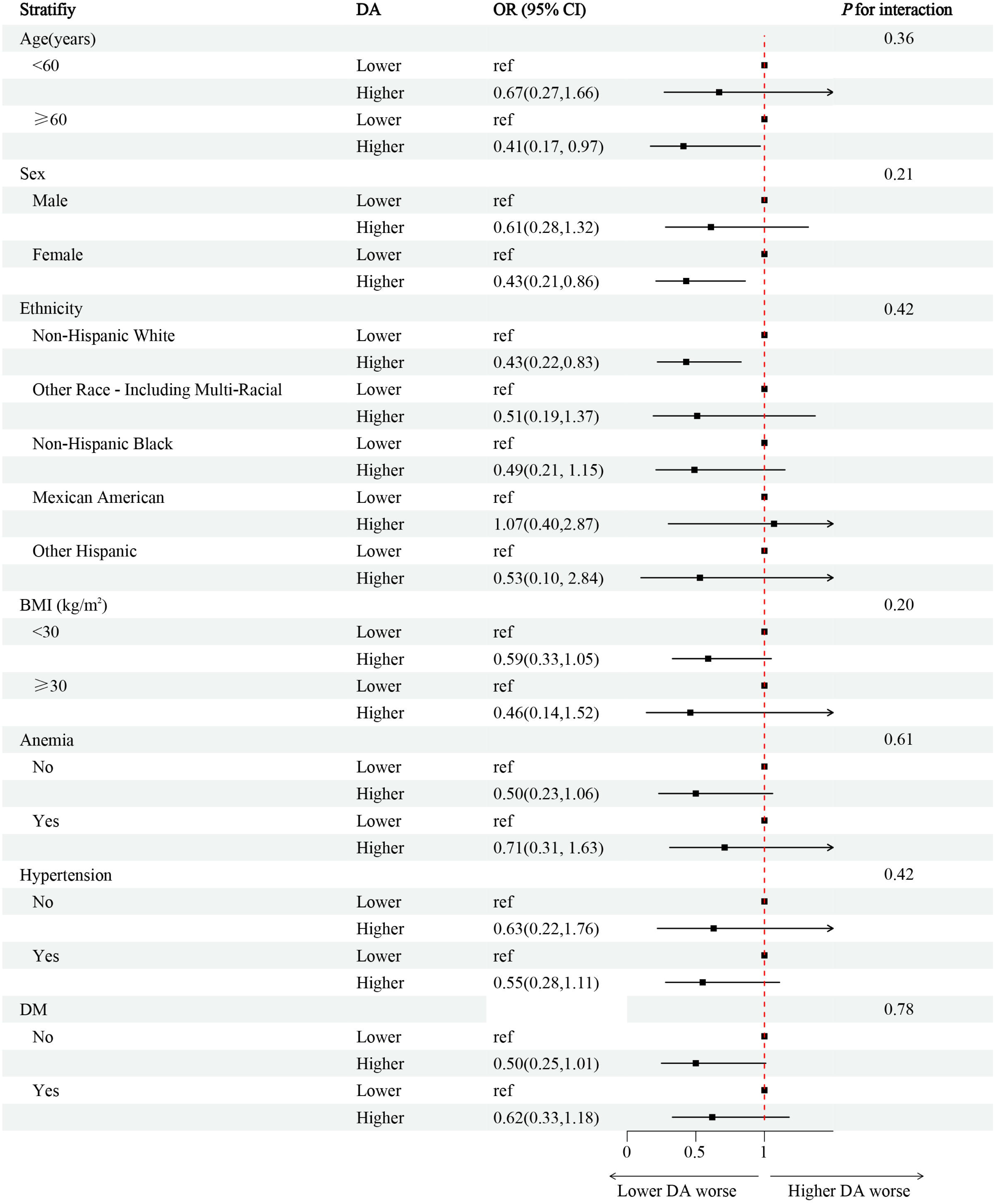

**Figure 5.**
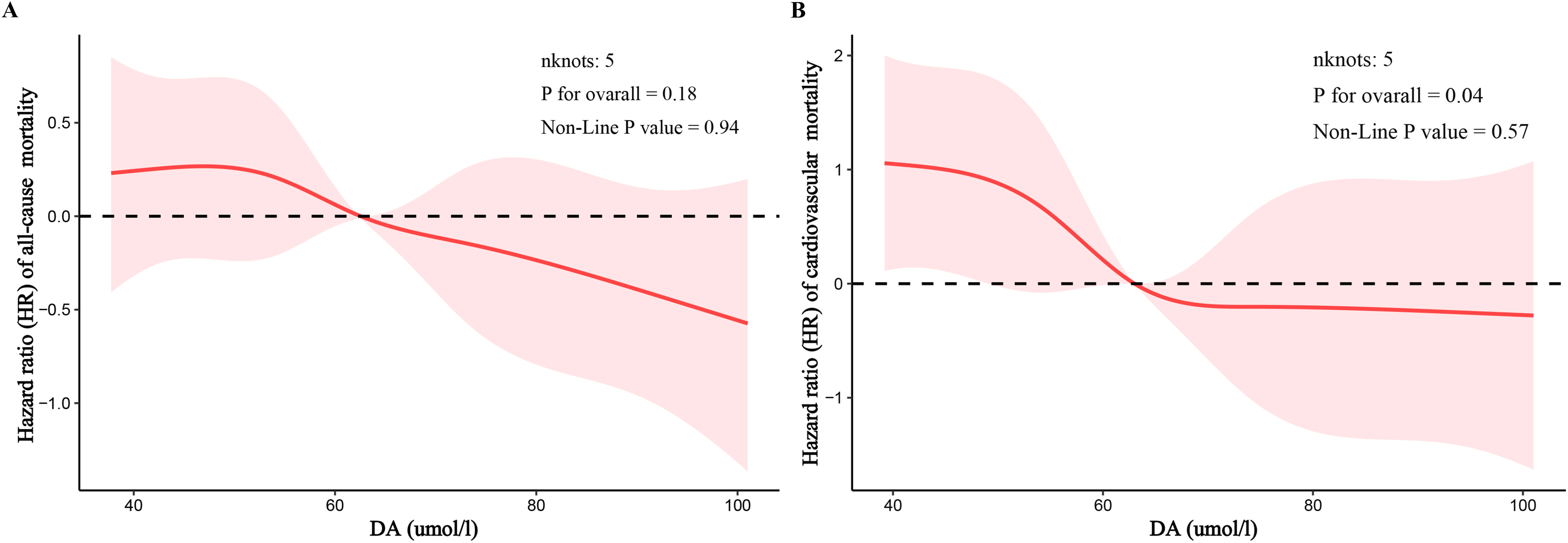

## 4. Discussion

This study found that the higher the circulating DA level, the lower the risk of CKD occurrence in the general population, and the lower the risk of cardiovascular and all-cause mortality in CKD patients.

DA, namely behenic acid, its metabolism mainly involves the synthesis and decomposition pathways of fatty acids, and is one of the very long-chain saturated fatty acids (VLCSFAs) ^16^. It can be endogenously produced in the human body, and its concentration can also be affected by dietary intake or lifestyle ^17^. The main source of DA in mammalian tissues is the endogenous synthesis achieved by elongation of saturated fatty acids (SFAs) with chain lengths of 12:0–18:0 (lauric acid to stearic acid) in the endoplasmic reticulum. This synthesis is mediated by a group of seven very long-chain fatty acid elongases (ELOVL 1–7) embedded in the endoplasmic reticulum ^2^. Currently, DA has been proved to be a biomarker of dietary intake and metabolic health, and is related to a series of health outcomes ^18,19^.

Currently study showed that DA was related to the occurrence of various diseases. Forouhi NG et al. ^19^ found that DA were negatively correlated with the risk of type 2 diabetes. Besides, DA is also associated with the occurrence of diseases such as atherosclerosis ^20^ and metabolic syndrome ^21^. Chiu YH et al. ^22^ found that there was a negative correlation between VLCSFA (including DA) on red blood cell membranes and the incidence rate of non-Hodgkin’s lymphoma except for chronic lymphocytic leukemia. Chung T-C et al. ^23^ proposed that DA could also be used as a potential biomarker for diagnosing hereditary diseases (such as X-linked adrenoleukodystrophy). Additionally, current studies have indicated that DA levels are negatively correlated with the occurrence risk of certain heart diseases, such as heart failure ^24^, cardiac arrest ^25^, atrial fibrillation ^26^, etc. Our research results show that there is a significant correlation between DA levels and the risk of CKD.

Furthermore, studies have found that the circulating DA was associated with a lower risk of unhealthy aging events ^27^. Liu M et al. ^28^ showed that a higher percentage of VLCSFAs (including DA) is related to positive cardiovascular health outcomes. Additionally, Fretts AM et al. ^29^ found in a large prospective study of 3,941 elderly people in the United States that the DA level was negatively correlated with the total mortality rate, with the risk of mortality reduced by 15%. A recent study also showed that high-level circulating DA has a protective effect on cardiovascular diseases, coronary heart disease and all-cause mortality in the entire population, the hyperlipidemia population and the hypertensive population ^7^. Our study resulted in similar conclusions. In CKD patients, the higher the DA level, the lower the risk of cardiovascular and all-cause mortality.

However, why is there a close association between DA levels and the occurrence and mortality of CKD? On the one hand, DA may reduce the risk of chronic kidney disease by improving the stability and function of cell membranes and alleviating the damage and stress of kidney cells. Ibarguren M et al. ^30^ found that by regulating fatty acids, the structure of membranes can be regulated, affecting cell processes dependent on this structure, and it is possible to reverse pathological cell dysfunctions that may trigger cancer, diabetes, hypertension, Alzheimer’s disease and Parkinson’s disease. Ardisson Korat AV et al. ^31^ found that the DA concentration in plasma and on red blood cell membranes was negatively correlated with the risk of T2D. It indicates that DA might provide certain protection to body cells by participating in the composition of cell membranes. On the other hand, DA is a component of ceramides, and the ceramide pathway has been proven to be crucial in L-homocysteine-induced glomerular damage and glomerulosclerosis ^9, 32^. In addition, DA may be involved in certain anti-inflammatory and antioxidant processes. DA undergoes β-oxidation in peroxisomes because mitochondria lack very long-chain acyl-CoA synthetase. The generated medium-chain acyl-CoA β-oxidation products can serve as precursors for plasmalogen synthesis in peroxisomes and undergo further modification in the endoplasmic reticulum ^33^. Plasmalogens are glycerophospholipid components of cell membranes and can be preferentially oxidized to prevent the oxidation of polyunsaturated fatty acids and other membrane components, thereby preventing the occurrence of lipid peroxidation ^8^, endowing cells with strong antioxidant functions, which may thereby reduce oxidative damage in cells. In addition to the antioxidant effect of DA’s own fatty acid metabolism, the unique regulatory effect of the body on DAβ oxidation also has an antioxidant effect. DA can be regulated by peroxisome proliferator-activated receptor (PPAR) δ, thereby regulating energy balance ^34^ and stimulating β-oxidation, triglyceride and glucose utilization in adipose tissue ^35^. Not only that, DA can also play a role in inflammatory responses ^9^. DA belongs to VLCFA and exhibits certain functions, such as functions in skin barrier formation, retinal function, myelin maintenance, sperm development and maturation, liver homeostasis regulation, and anti-inflammation^2^. These functions are not caused by free VLCFAs themselves, but are caused by their influence as components of cell membrane lipids (sphingolipids and glycerophospholipids) or as precursors of lipid mediators that can alleviate inflammation ^2^. Activation of the DA receptor PPAR δ can reduce the production of pro-inflammatory cytokines involved in insulin resistance and improve the function of pancreatic B cells ^36^. In addition, this receptor plays an important role in controlling lipid metabolism, atherosclerosis and anti-inflammation ^37^. To sum up, the association between DA and the risk of occurrence and mortality of CKD may be attributed to the physiological functions of DA, its antioxidant stress and anti-inflammatory effects, and its protective effects on cardiovascular diseases, among other multi-faceted effects.

However, our study also has certain limitations. First, this study only preliminarily revealed the association between DA levels and the risk of CKD occurrence and mortality, and the causal relationship and specific mechanism of action still need to be further explored in depth. Secondly, due to the relatively limited sample size, it may have a certain impact on the universality of the results. In addition, other potential confounding factors may not be completely controlled. Finally, when exploring the association between DA levels and the risk of cardiovascular and all-cause mortality in CKD patients, because the included sample size was small, no grouping processing, subgroup analysis, and sensitivity analysis were performed.

In conclusion, our research results indicate that the higher the DA level, the lower the risk of CKD occurrence in individuals; and the higher the DA level, the lower the risk of mortality in CKD patients. Clinicians should pay attention to the individual circulating DA level, which may help screen individuals with a high risk of CKD in the general population and patients with a high risk of mortality among CKD patients.

## Funding

The Science and technology fund of Chengdu Medical College (CYZYB22-02).

The research fund of Sichuan Medical and Health Care Promotion institute (KY2022QN0309).

The Sichuan Provincial Medical Association Youth Innovation Project (Q23021).

The Sichuan Province Science and Technology Project (No. 2023NSFSC1529).

## Author Contributions

Conception and design of the study: Xiang Xiao, JunLin Zhang, Xianglong Meng, Qianyu Yang, Zhuoxing Li. Acquisition and analysis of data: Xiang Xiao, JunLin Zhang,Xianglong Meng, Qianyu Yang; Drafting the manuscript or figures: Xiang Xiao, Qianyu Yang, JunLin Zhang, Xianglong Meng, Wan Li, Ping Zhou, Wuyu Gao, Tingyu Wu, Haiyan Yu, Guifei Deng, Qiyuan Liang.

## Data Availability

Some or all datasets generated during and/or analyzed during the current study are not publicly available but are available from the corresponding author on reasonable request.

## Acknowledgments

The authors wish to thank all the participants of this study for their important contributions.

## Ethical Approval

The study protocol conformed to the ethical standards of the 1964 Declaration of Helsinki and its subsequent amendments, with approval from the National Committee for Ethical Review of Health Statistics Research and signed informed consent from all participants.

## Declarations

The authors declare that they have no known competing financial interests or personal relationships that could have appeared to influence the work reported in this paper.

