## Supplemental Table 1,2,3,4,5,6,7,8,9 for "The association between docosanoic acid and the risks of occurrence and mortality of chronic kidney disease": Supplementary_DA_CKD_siwang.docx

**Supplementary -table 1. Definitions/criteria of some diagnoses**

| Variables | Definitions/criteria |
| --- | --- |
| Smoker | Smoking more than 100 cigarettes in previous and now. |
| Alcohol user ^1^ | ≥2 drinks per day for females, ≥3 drinks per day for males, or binge drinking ≥2 days per month.  Binge drinking (≥4 drinks on the same occasion for females, ≥5 drinks on the same occasion for males) on 5 or more days per month. |
| Hypertension ^2^ | (1) Self-reported hypertension diagnosis, (2) Use of anti-hypertensive medication, (3) Average systolic blood pressure (SBP) > 140 mmHg, (4) Average diastolic blood pressure (DBP) > 90 mmHg, meet any of the above conditions. |
| Anemia ^3^ | ≥120g/L for women (15 years of age and above), ≥130g/L for men (15 years of age and above). |
| Hyperlipidemia | (1) Triglyceridemia ≥ 150 mg/dl; (2) Hypercholesterolemia: a) total cholesterol ≥ 200 mg/dl, b) low-density lipoprotein ≥ 130 mg/dl), c). high-density lipoprotein (< 40 mg/dl, male; < 50 mg/dl, female), meet any of the above conditions; (3) Use of lipid-lowering drugs; meet any of the above conditions. |
| Diabetes mellitus | 1) a doctor has told you that you have diabetes, 2) HbA1c (%) > 6.5, 3) fasting blood glucose (mmol/l) ≥ 7.0, 4) random blood glucose (mmol/l) ≥ 11.1, and 5) two-hour oral glucose tolerance test (OGTT) blood glucose (mmol/l) >= 11.1 ^4^. |

**Supplementary -table 2. Logistic-regression analysis of risk factors for CKD in individuals**

| Variables | Unadjusted | | Model 1 | | Model 2 | | Model 3 | |
| --- | --- | --- | --- | --- | --- | --- | --- | --- |
|  | OR (95% CI) | *P*-value |  | *P*-value |  | *P*-value | OR (95% CI) | *P*-value |
| DA |  |  |  |  |  |  |  |  |
| Lower | ref |  | ref |  | ref |  | ref |  |
| Higher | 0.61(0.47,0.80) | 0.001 | 0.50(0.36,0.70) | 0.001 | 0.50(0.26,0.98) | 0.04 | 0.58(0.39,0.87) | 0.01 |
| Age | 1.06(1.04,1.08) | <0.001 | 1.06(1.04,1.08) | <0.001 | 1.06(1.03,1.09) | 0.01 | 1.04(1.02,1.06) | <0.001 |
| Sex |  |  |  |  |  |  |  |  |
| Female | ref |  | ref |  | ref |  | ref |  |
| Male | 0.76(0.55,1.05) | 0.09 | 0.70(0.44,1.10) | 0.11 | 0.61(0.36,1.04) | 0.06 | 0.62(0.44,0.87) | 0.01 |
| Ethnicity |  |  |  |  |  |  |  |  |
| Mexican American | ref |  | 0.89(0.52,1.50) | 0.61 | 0.82(0.28,2.40) | 0.6 | ref |  |
| Non-Hispanic Black | 1.14(0.69,1.88) | 0.59 | 0.51(0.33,0.80) | 0.01 | 0.59(0.19,1.88) | 0.24 | 0.61(0.32,1.17) | 0.13 |
| Non-Hispanic White | 0.87(0.55,1.39) | 0.54 | 0.55(0.35,0.88) | 0.02 | 0.56(0.25,1.27) | 0.11 | 0.61(0.30,1.25) | 0.16 |
| Other Hispanic | 0.76(0.45,1.28) | 0.27 | 0.91(0.58,1.42) | 0.64 | 1.02(0.54,1.94) | 0.92 | 0.60(0.37,0.98) | 0.04 |
| Other Race - Including Multi-Racial | 1.10(0.71,1.69) | 0.65 | 0.89(0.52,1.50) | 0.61 | 0.82(0.28,2.40) | 0.60 | 0.88(0.55,1.43) | 0.59 |
| BMI (kg/m2) |  |  |  |  |  |  |  |  |
| <30 | ref |  | ref |  | ref |  | ref |  |
| ≥30 | 1.76(1.31,2.37) | <0.001 | 1.66(1.22,2.26) | 0.005 | 1.70(0.94,3.05) | 0.06 | 1.19(0.79,1.80) | 0.39 |
| Alcohol use |  |  |  |  |  |  |  |  |
| No | ref |  |  |  | ref |  | ref |  |
| Yes | 0.49(0.34,0.71) | <0.001 |  |  | 0.82(0.44,1.54) | 0.39 | 0.77(0.49,1.22) | 0.25 |
| Smoke |  |  |  |  |  |  |  |  |
| No | ref |  |  |  | ref |  | ref |  |
| Yes | 1.51(1.02,2.23) | 0.04 |  |  | 1.19(0.52,2.73) | 0.56 | 1.19(0.66,2.13) | 0.54 |
| Poverty (dollar) (%) |  |  |  |  |  |  |  |  |
| 0-1 | ref |  |  |  | ref |  | ref |  |
| 1.1-3 | 1.80(1.11,2.94) | 0.02 |  |  | 2.02(1.04,3.93) | 0.04 | 1.76(1.10,2.81) | 0.02 |
| >3 | 1.80(1.11,2.94) | 0.004 |  |  | 1.43(0.87,2.37) | 0.11 | 1.43(1.00,2.03) | 0.05 |
| Education (%) |  |  |  |  |  |  |  |  |
| High school or equivalent | ref |  |  |  | ref |  | ref |  |
| Less than high school | 1.68(1.00,2.84) | 0.05 |  |  | 1.26(0.63,2.52) | 0.36 | 1.16(0.69,1.94) | 0.56 |
| College or above | 2.35(1.17,4.72) | 0.02 |  |  | 1.03(0.32,3.32) | 0.94 | 0.90(0.41,1.98) | 0.78 |
| Anemia |  |  |  |  |  |  |  |  |
| No | ref |  |  |  |  |  | ref |  |
| Yes | 3.70(2.44,5.61) | <0.001 |  |  |  |  | 2.28(1.28,4.08) | 0.01 |
| Hyperlipidemia |  |  |  |  |  |  |  |  |
| No | ref |  |  |  |  |  | ref |  |
| Yes | 2.14(1.26,3.62) | 0.01 |  |  |  |  | 1.04(0.59,1.82) | 0.90 |
| Hypertension |  |  |  |  |  |  |  |  |
| No | ref |  |  |  |  |  | ref |  |
| Yes | 4.55(3.14,6.61) | <0.001 |  |  |  |  | 1.92(1.31,2.82) | 0.002 |
| Diabetes mellitus |  |  |  |  |  |  |  |  |
| No | ref |  |  |  |  |  | ref |  |
| Yes | 5.63(4.38,7.24) | <0.001 |  |  |  |  | 2.37(1.47,3.83) | 0.001 |
| RAASi use |  |  |  |  |  |  |  |  |
| No | ref |  |  |  |  |  | ref |  |
| Yes | 3.48(2.69,4.50) | <0.001 |  |  |  |  | 0.99(0.66,1.49) | 0.97 |

**Model 1** adjusted for baseline age, sex, ethnicity, BMI; **Model 2** adjusted for covariates in model 1 plus smoke (‘yes’ or ‘no’), alcohol use (‘yes’ or ‘no’), poverty, education; **Model 3** adjusted for covariates in model 2 plus anemia (‘yes’ or ‘no’), hyperlipidemia (‘yes’ or ‘no’), hypertension (‘yes’ or ‘no’), DM (‘yes’ or ‘no’), RAASi use (‘yes’ or ‘no’). DA, docosanoic acid; CKD, chronic kidney disease; OR, Odd Ratio; CI, confidence interval; BMI, Body Mass Index; DM, diabetes mellitus; RAASi, renin-angiotensin system inhibitors.

**Supplementary -table 3. Multifactor logistic regression analysis of risk factors for the risk of** **CKD in individuals**

| Variables | Unadjusted | | Model 1**^a^** | | Model 2**^b^** | | Model 3**^c^** | |
| --- | --- | --- | --- | --- | --- | --- | --- | --- |
|  | OR (95% CI) | *P*-value | OR (95% CI) | P-value | OR (95% CI) | *P*-value | OR (95% CI) | *P*-value |
| Lower | ref |  | ref |  | ref |  | ref | ref |
| Higher | 0.61(0.47,0.80) | 0.001 | 0.50(0.36,0.70) | 0.001 | 0.50(0.26,0.98) | 0.04 | 0.58(0.39,0.87) | 0.01 |
| Per-SD increment of DA | 0.81(0.70,0.93) | 0.01 | 0.73(0.63,0.85) | 0.001 | 0.74(0.57,0.98) | 0.04 | 0.82(0.70,0.96) | 0.02 |

**Model 1** adjusted for baseline age, sex, ethnicity, BMI; **Model 2** adjusted for covariates in model 1 plus smoke (‘yes’ or ‘no’), alcohol use (‘yes’ or ‘no’), poverty, education; **Model 3** adjusted for covariates in model 2 plus anemia (‘yes’ or ‘no’), hyperlipidemia (‘yes’ or ‘no’), hypertension (‘yes’ or ‘no’), DM (‘yes’ or ‘no’), RAASi use (‘yes’ or ‘no’). DA, docosanoic acid; CKD, chronic kidney disease; OR, Odd Ratio; CI, confidence interval; BMI, Body Mass Index; DM, diabetes mellitus; RAASi, renin-angiotensin system inhibitors.

**Supplementary -table 4. Stratified analysis of DA and the risk of CKD in individuals**

| Variable | OR（95% CI） | | *P* for interaction |
| --- | --- | --- | --- |
|  | Lower | Higher |  |
| Age (years) |  |  | 0.36 |
| <60 | ref | 0.67(0.27,1.66) |  |
| ≥60 | ref | 0.41(0.17, 0.97) |  |
| Sex |  |  | 0.21 |
| Male | ref | 0.61(0.28,1.32) |  |
| Female | ref | 0.43(0.21,0.86) |  |
| Ethnicity |  |  | 0.42 |
| Non-Hispanic White | ref | 0.43(0.22,0.83) |  |
| Other Race - Including Multi-Racial | ref | 0.51(0.19,1.37) |  |
| Non-Hispanic Black | ref | 0.49(0.21, 1.15) |  |
| Mexican American | ref | 1.07(0.40,2.87) |  |
| Other Hispanic | ref | 0.53(0.10, 2.84) |  |
| BMI (kg/m^2^) |  |  | 0.20 |
| <30 | ref | 0.59(0.33,1.05) |  |
| ≥30 | ref | 0.46(0.14,1.52) |  |
| Hypertension |  |  | 0.91 |
| No | ref | 0.63(0.22,1.76) |  |
| Yes | ref | 0.55(0.28,1.11) |  |
| DM |  |  | 0.78 |
| No | ref | 0.50(0.25,1.01) |  |
| Yes | ref | 0.62(0.33,1.18) |  |

DA, docosanoic acid; OR, odds ratio; CI, Confidence interval; BMI, body mass index; DM, diabetes mellitus.

**Supplementary -table 5. The relationship between DA and the risk of CKD based on the data after multiple imputation.**

| Variables | Unadjusted | | Model 1 | | Model 2 | | Model 3 | |
| --- | --- | --- | --- | --- | --- | --- | --- | --- |
|  | OR (95% CI) | *P*-value | OR (95% CI) | P-value | OR (95% CI) | *P*-value | OR (95% CI) | *P*-value |
| Lower | ref |  | ref |  | ref |  | ref | ref |
| Higher | 0.61(0.47,0.80) | 0.001 | 0.50(0.36,0.70) | 0.001 | 0.54(0.29,0.98) | 0.05 | 0.61(0.43,0.88) | 0.01 |

**Model 1** adjusted for baseline age, sex, ethnicity, BMI; **Model 2** adjusted for covariates in model 1 plus smoke (‘yes’ or ‘no’), alcohol use (‘yes’ or ‘no’), poverty, education; **Model 3** adjusted for covariates in model 2 plus anemia (‘yes’ or ‘no’), hyperlipidemia (‘yes’ or ‘no’), hypertension (‘yes’ or ‘no’), DM (‘yes’ or ‘no’), RAASi use (‘yes’ or ‘no’). DA, docosanoic acid; CKD, chronic kidney disease; OR, Odd Ratio; CI, confidence interval; BMI, Body Mass Index; DM, diabetes mellitus; RAASi, renin-angiotensin system inhibitors.

**Supplementary -table 6. Associations between DA and the risk of** **CKD in individuals as determined by tertiles grouping of DA**

| Variables | Unadjusted | | Model 1 | | Model 2 | | Model 3 | |
| --- | --- | --- | --- | --- | --- | --- | --- | --- |
|  | OR (95% CI) | *P*-value | OR (95% CI) | P-value | OR (95% CI) | *P*-value | OR (95% CI) | *P*-value |
| T1 | ref |  | ref |  | ref |  | ref | ref |
| T2 | 0.98(0.72,1.33) | 0.88 | 0.87(0.58,1.30) | 0.44 | 0.89(0.37,2.15) | 0.62 | 1.07(0.69,1.68) | 0.74 |
| T3 | 0.67(0.48,0.93) | 0.02 | 0.53(0.36,0.77) | 0.005 | 0.59(0.39,0.88) | 0.01 | 0.64(0.43,0.95) | 0.03 |
| *P* for trend |  | 0.02 |  | 0.004 |  | 0.04 |  | 0.02 |

**Model 1** adjusted for baseline age, sex, ethnicity, BMI; **Model 2** adjusted for covariates in model 1 plus smoke (‘yes’ or ‘no’), alcohol use (‘yes’ or ‘no’), poverty, education; **Model 3** adjusted for covariates in model 2 plus anemia (‘yes’ or ‘no’), hyperlipidemia (‘yes’ or ‘no’), hypertension (‘yes’ or ‘no’), DM (‘yes’ or ‘no’), RAASi use (‘yes’ or ‘no’). DA, docosanoic acid; CKD, chronic kidney disease; OR, Odd Ratio; CI, confidence interval; BMI, Body Mass Index; DM, diabetes mellitus; RAASi, renin-angiotensin system inhibitors.

**Supplementary -table 7. Baseline clinical features of enrolled individuals**

| Variable | Total (N=2366) | Without CKD  (n_1_=1958) | CKD  (n_2_=408) | *P*-value |
| --- | --- | --- | --- | --- |
| DA (umol/L) | 68.56(0.80) | 69.02(0.87) | 65.66(0.86) | 0.01 |
| Age (years) | 48.05(0.67) | 46.08(0.69) | 60.52(1.51) | < 0.001 |
| Sex (%) |  |  |  | 0.09 |
| Female | 50.99(0.04) | 50.06(1.57) | 56.84(3.67) |  |
| Male | 49.01(0.04) | 49.94(1.57) | 43.16(3.67) |  |
| Ethnicity (%) |  |  |  | 0.002 |
| Mexican American | 7.86(0.02) | 10.19(2.13) | 5.87(1.40) |  |
| Non-Hispanic Black | 9.35(0.01) | 8.79(1.74) | 9.83(1.95) |  |
| Non-Hispanic White | 68.77(0.08) | 64.24(3.32) | 72.64(3.78) |  |
| Other Hispanic | 6.52(0.01) | 7.98(1.58) | 5.27(1.61) |  |
| Other Race - Including Multi-Racial | 7.51(0.01) | 8.80(1.51) | 6.40(1.35) |  |
| BMI (kg/m^2^) (%) |  |  |  | < 0.001 |
| <30 | 63.39(0.05) | 65.66(1.80) | 52.06(3.73) |  |
| ≥30 | 35.94(0.03) | 34.34(1.80) | 47.94(3.73) |  |
| Alcohol use (%) |  |  |  | < 0.001 |
| No | 9.07(0.01) | 8.90(0.65) | 16.62(2.58) |  |
| Yes | 81.67(0.07) | 91.10(0.65) | 83.38(2.58) |  |
| Smoke (%) |  |  |  | 0.04 |
| No | 56.01(0.04) | 57.43(1.28) | 47.26(5.02) |  |
| Yes | 43.93(0.04) | 42.57(1.28) | 52.74(5.02) |  |
| Serum albumin (g/L) | 42.97(0.09) | 43.11(0.11) | 42.07(0.24) | 0.002 |
| ACR (mg/g) | 29.17(3.00) | 7.75( 0.13) | 167.65(20.63) | < 0.001 |
| ACR (mg/g) |  |  |  | < 0.001 |
| <30 | 89.87(0.07) | 100.00(0.00) | 26.59(4.05) |  |
| (30-300) | 8.36(0.01) | 0.00(0.00) | 62.64(3.36) |  |
| ≥300 | 1.44(0.00) | 0.00(0.00) | 10.76(1.76) |  |
| Anemia(%) |  |  |  | < 0.001 |
| 0 | 93.22(0.07) | 94.81(0.63) | 83.16(1.68) |  |
| 1 | 6.78(0.01) | 5.19(0.63) | 16.84(1.68) |  |
| Hypertension(%) |  |  |  | < 0.001 |
| No | 60.77(0.05) | 65.71(2.12) | 29.62(4.00) |  |
| Yes | 39.23(0.04) | 34.29(2.12) | 70.38(4.00) |  |
| DM |  |  |  | < 0.001 |
| NO | 83.88(0.06) | 88.15(1.49) | 56.93(3.29) |  |
| Yes | 16.12(0.02) | 11.85(1.49) | 43.07(3.29) |  |
| Hyperlipidemia (%) |  |  |  | 0.01 |
| No | 26.61(0.02) | 28.35(1.15) | 15.61(3.51) |  |
| Yes | 73.39(0.06) | 71.65(1.15) | 84.39(3.51) |  |
| eGFR (ml/min/1.73m^2^) | 94.88(0.78) | 97.94(0.56) | 75.38(3.18) | < 0.001 |
| eGFR (ml/min/1.73m^2^) (%) |  |  |  | < 0.001 |
| >60 | 93.77(0.07) | 100.00(0.00) | 55.01(5.47) |  |
| <60 | 6.09(0.01) | 0.00(0.00) | 44.99(5.47) |  |
| RAASi use (%) |  |  |  | < 0.001 |
| No | 80.94(0.06) | 84.24(1.45) | 60.59(3.81) |  |
| Yes | 18.96(0.02) | 15.76(1.45) | 39.41(3.81) |  |
| Mortality |  |  |  | < 0.001 |
| No | 91.42(0.07) | 95.00(0.59) | 69.12(2.43) |  |
| Yes | 8.54(0.01) | 5.00(0.59) | 30.88(2.43) |  |

DA, docosanoic acid; BMI, Body Mass Index; ACR, albumin-to-creatinine ratio; DM, diabetes mellitus; eGFR, estimated glomerular filtration rate; RAASi, renin-angiotensin system inhibitors.

**Supplementary -table 8. Association between DA and risk of cardiovascular mortality in patients with CKD**

| Variables | Unadjusted | | Model 1 | | Model 2 | | Model 3 | |
| --- | --- | --- | --- | --- | --- | --- | --- | --- |
|  | HR (95% CI) | *P*-value | HR (95% CI) | *P*-value | HR (95% CI) | *P*-value | HR (95% CI) | *P*-value |
| Per-SD increment of DA | 0.60(0.36,1.00) | **0.05** | 0.55(0.36,0.85) | **0.01** | 0.55(0.35,0.85) | **0.01** | 0.56(0.35,0.90) | **0.02** |
| Age | 1.11(1.08,1.14) | **<0.0001** | 1.11(1.08,1.14) | **<0.0001** | 1.10(1.07,1.13) | **<0.0001** | 1.11(1.05,1.16) | **<0.0001** |
| Sex |  |  |  |  |  |  |  |  |
| Female | ref |  | ref |  | ref |  | ref |  |
| Male | 0.78(0.32,1.92) | 0.59 | 0.64(0.30,1.36) | 0.25 | 0.71(0.38,1.32) | 0.28 | 0.66(0.34,1.25) | 0.2 |
| Ethnicity |  |  |  |  |  |  |  |  |
| Mexican American | ref |  | ref |  | ref |  | ref |  |
| Non-Hispanic Black | 1.10(0.36,3.35) | 0.87 | 0.51(0.22,1.18) | 0.11 | 0.47(0.20,1.12) | 0.09 | 0.47(0.17,1.30) | 0.14 |
| Non-Hispanic White | 1.68(0.66,4.27) | 0.28 | 0.43(0.18,1.02) | **0.05** | 0.47(0.16,1.37) | 0.17 | 0.57(0.20,1.64) | 0.3 |
| Other Hispanic | 1.51(0.36,6.30) | 0.57 | 0.53(0.16,1.79) | 0.31 | 0.71(0.23,2.25) | 0.57 | 1.42(0.32,6.38) | 0.64 |
| Other Race - Including Multi-Racial | 0.95(0.09,9.58) | 0.96 | 0.27(0.05,1.54) | 0.14 | 0.26(0.05,1.43) | 0.12 | 0.25(0.04,1.61) | 0.15 |
| BMI (kg/m^2^) |  |  |  |  |  |  |  |  |
| <30 | ref |  | ref |  | ref |  | ref |  |
| ≥30 | 0.92(0.52,1.62) | 0.77 | 0.78(0.49,1.25) | 0.3 | 0.74(0.46,1.20) | 0.22 | 0.56(0.28,1.13) | 0.1 |
| Alcohol use |  |  |  |  |  |  |  |  |
| No | ref |  |  |  | ref |  | ref |  |
| Yes | 0.42(0.17,1.03) | 0.06 |  |  | 0.72(0.29,1.77) | 0.48 | 0.53(0.23,1.22) | 0.14 |
| Smoke |  |  |  |  |  |  |  |  |
| No | ref |  |  |  | ref |  | ref |  |
| Yes | 0.93(0.43,2.00) | **0.85** |  |  | 0.99(0.49,2.01) | 0.97 | 1.62(0.69,3.83) | 0.27 |
| Anemia |  |  |  |  |  |  |  |  |
| No | ref |  |  |  |  |  | ref |  |
| Yes | 1.41(0.64,3.12) | 0.39 |  |  |  |  | 0.65(0.20,2.11) | 0.48 |
| Hyperlipidemia |  |  |  |  |  |  |  |  |
| No | ref |  |  |  |  |  | ref |  |
| Yes | 1.76(0.52,5.97) | 0.36 |  |  |  |  | 1.15(0.26,5.19) | 0.85 |
| Hypertension |  |  |  |  |  |  |  |  |
| No | ref |  |  |  |  |  | ref |  |
| Yes | 2.94(0.99,8.76) | **0.05** |  |  |  |  | 0.66(0.22,1.92) | 0.44 |
| DM |  |  |  |  |  |  |  |  |
| No | ref |  |  |  |  |  | ref |  |
| Yes | 3.52(1.71,7.26) | <0.001 |  |  |  |  | 2.37(0.85,6.61) | 0.10 |
| RAASi use |  |  |  |  |  |  |  |  |
| No | ref |  |  |  |  |  | ref |  |
| Yes | 1.64(0.72,3.78) | 0.24 |  |  |  |  | 0.98(0.43,2.23) | 0.96 |
| ACR | 1.00(1.00,1.00) | **<0.001** |  |  |  |  | 1.00(1.00,1.00) | **0.001** |
| eGFR | 0.98(0.96,0.99) | **<0.001** |  |  |  |  | 1.01(0.98,1.04) | 0.64 |

**Model 1** adjusted for baseline age, sex, ethnicity, BMI; **Model 2** adjusted for covariates in model 1 plus smoke (‘yes’ or ‘no’), alcohol use (‘yes’ or ‘no’); **Model 3** adjusted for covariates in model 2 plus anemia (‘yes’ or ‘no’), hyperlipidemia (‘yes’ or ‘no’), hypertension (‘yes’ or ‘no’), DM (‘yes’ or ‘no’), RAASi use (‘yes’ or ‘no’), ACR, eGFR. DA, docosanoic acid; CKD, chronic kidney disease; HR, Hazard Ratio; CI, confidence interval; BMI, Body Mass Index; DM, diabetes mellitus; RAASi, renin-angiotensin system inhibitors; ACR, Albumin Creatinine Ratio; eGFR, Estimated Glomerular Filtration Rate.

**Supplementary -table 9. Association between DA and risk of all-cause mortality in patients with CKD**

| Variables | Unadjusted | | Model 1 | | Model 2 | | Model 3 | |
| --- | --- | --- | --- | --- | --- | --- | --- | --- |
|  | HR (95% CI) | P-value | HR (95% CI) | P-value | HR (95% CI) | P-value | HR (95% CI) | P-value |
| Per-SD increment of DA | 0.78(0.62,0.99) | 0.78 | 0.77(0.65,0.91) | 0.002 | 0.76(0.64,0.91) | 0.002 | 0.73(0.59,0.89) | 0.002 |
| Age | 1.06(1.04,1.08) | <0.001 | 1.08(1.05,1.10) | <0.001 | 1.07(1.04,1.10) | <0.001 | 1.08(1.06,1.10) | <0.0001 |
| Sex |  |  |  |  |  |  |  |  |
| Female | ref |  | ref |  | ref |  | ref |  |
| Male | 1.11(0.58,2.11) | 0.75 | 1.09(0.59,1.99) | 0.79 | 1.03(0.58,1.84) | 0.91 | 0.93(0.55,1.59) | 0.8 |
| Ethnicity |  |  |  |  |  |  |  |  |
| Mexican American | ref |  |  |  | ref |  | ref |  |
| Non-Hispanic Black | 1.93(1.15,3.23) | 0.01 | 1.05(0.60,1.86) | 0.86 | 1.08(0.59,1.97) | 0.8 | 1.19(0.58,2.43) | 0.64 |
| Non-Hispanic White | 3.26(1.73,6.13) | <0.001 | 1.07(0.52,2.19) | 0.85 | 1.08(0.49,2.40) | 0.85 | 1.59(0.69,3.71) | 0.28 |
| Other Hispanic | 3.13(1.34,7.30) | 0.01 | 1.35(0.71,2.58) | 0.36 | 1.72(0.90,3.28) | 0.1 | 2.64(1.22,5.70) | 0.01 |
| Other Race - Including Multi-Racial | 1.81(0.73,4.47) | 0.2 | 0.66(0.28,1.52) | 0.33 | 0.66(0.28,1.56) | 0.34 | 0.92(0.35,2.45) | 0.87 |
| BMI (kg/m2) |  |  |  |  |  |  |  |  |
| <30 | ref |  | ref |  | ref |  | ref |  |
| ≥30 | 1.00(0.67,1.50) | 0.99 | 1.00(0.68,1.48) | 0.99 | 1.06(0.75,1.52) | 0.73 | 1.21(0.68,2.15) | 0.52 |
| Alcohol use |  |  |  |  |  |  |  |  |
| No | ref |  |  |  | ref |  | ref |  |
| Yes | 0.76(0.41,1.39) | 0.37 |  |  | 0.89(0.45,1.75) | 0.73 | 0.76(0.35,1.64) | 0.48 |
| Smoke |  |  |  |  |  |  |  |  |
| No | ref |  |  |  | ref |  | ref |  |
| Yes | 1.59(1.00,2.52) | 0.05 |  |  | 1.44(0.89,2.32) | 0.13 | 1.77(1.02,3.09) | 0.04 |
| Anemia |  |  |  |  |  |  |  |  |
| No | ref |  |  |  |  |  | ref |  |
| Yes | 1.38(0.82,2.34) | 0,23 |  |  |  |  | 1.12(0.61,2.04) | 0.72 |
| Hyperlipidemia |  |  |  |  |  |  |  |  |
| No | ref |  |  |  |  |  | ref |  |
| Yes | 1.64(0.80,3.37) | 0.17 |  |  |  |  | 1.67(0.90,3.11) | 0.10 |
| Hypertension |  |  |  |  |  |  |  |  |
| No | ref |  |  |  |  |  | ref |  |
| Yes | 2.28(1.33,3.90) | <0.003 |  |  |  |  | 0.63(0.28,1.40) | 0.25 |
| Diabetes mellitus |  |  |  |  |  |  |  |  |
| No | ref |  |  |  |  |  | ref |  |
| Yes | 1.80(1.13,2.85) | 0.01 |  |  |  |  | 1.19(0.61,2.33) | 0.62 |
| RAASi use |  |  |  |  |  |  |  |  |
| No | ref |  |  |  |  |  | ref |  |
| Yes | 1.50(0.93,2.42) | 0.1 |  |  |  |  | 0.85(0.46,1.55) | 0.59 |
| ACR | 1.00(1.00,1.00) | <0.001 |  |  |  |  | 1.00(1.00,1.00) | <0.001 |
| eGFR | 0.98(0.97,0.99) | <0.001 |  |  |  |  | 1.01(1.00,1.02) | 0.26 |

**Model 1** adjusted for baseline age, sex, ethnicity, BMI; **Model 2** adjusted for covariates in model 1 plus smoke (‘yes’ or ‘no’), alcohol use (‘yes’ or ‘no’); **Model 3** adjusted for covariates in model 2 plus anemia (‘yes’ or ‘no’), hyperlipidemia (‘yes’ or ‘no’), hypertension (‘yes’ or ‘no’), DM (‘yes’ or ‘no’), RAASi use (‘yes’ or ‘no’), ACR, eGFR. DA, docosanoic acid; CKD, chronic kidney disease; HR, Hazard Ratio; CI, confidence interval; BMI, Body Mass Index; DM, diabetes mellitus; RAASi, renin-angiotensin system inhibitors; ACR, Albumin Creatinine Ratio; eGFR, Estimated Glomerular Filtration Rate.
