## Supplementary material for "The association between docosanoic acid and the risks of occurrence and mortality of chronic kidney disease": Table 1,2: table_DA_CKD_siwang.docx

**Table 1. Baseline clinical features of enrolled individuals**

| Variable | Total (N=2366) | Lower DA  (≤65.10)  (n_1_=1185) | Higher DA  (＞65.10)  (n_2_=1181) | *P*-value |
| --- | --- | --- | --- | --- |
| DA (umol/L) | 68.56(0.80) | 54.80(0.27) | 80.29(0.66) | < 0.001 |
| Age (years) | 48.05(0.67) | 46.20(0.83) | 49.63(0.72) | 0.001 |
| Sex (%) |  |  |  | < 0.001 |
| Female | 50.99(0.04) | 43.17(2.13) | 57.66(1.84) |  |
| Male | 49.01(0.04) | 56.83(2.13) | 42.34(1.84) |  |
| Ethnicity (%) |  |  |  | 0.002 |
| Mexican American | 7.86(0.02) | 10.19(2.13) | 5.87(1.40) |  |
| Non-Hispanic Black | 9.35(0.01) | 8.79(1.74) | 9.83(1.95) |  |
| Non-Hispanic White | 68.77(0.08) | 64.24(3.32) | 72.64(3.78) |  |
| Other Hispanic | 6.52(0.01) | 7.98(1.58) | 5.27(1.61) |  |
| Other Race - Including Multi-Racial | 7.51(0.01) | 8.80(1.51) | 6.40(1.35) |  |
| BMI (kg/m^2^) (%) |  |  |  | 0.45 |
| <30 | 63.39(0.05) | 64.87(1.46) | 62.92(2.75) |  |
| ≥30 | 35.94(0.03) | 35.13(1.46) | 37.08(2.75) |  |
| Alcohol use (%) |  |  |  | 0.27 |
| No | 9.07(0.01) | 9.15(1.09) | 10.69(0.96) |  |
| Yes | 81.67(0.07) | 90.85(1.09) | 89.31(0.96) |  |
| Poverty (dollar) (%) |  |  |  | 0.04 |
| 0-1 | 15.48(0.02) | 19.12(1.67) | 14.42(2.16) |  |
| 1.1-3 | 33.79(0.04) | 37.27(3.27) | 35.28(2.95) |  |
| >3 | 44.08(0.05) | 43.62(3.52) | 50.30(4.45) |  |
| Education (%) |  |  |  | 0.04 |
| High school or equivalent | 31.04(0.04) | 32.96(2.46) | 29.41(3.05) |  |
| Less than high school | 6.34(0.01) | 7.93(1.15) | 4.99(0.66) |  |
| College or above | 62.61(0.05) | 59.11(2.69) | 65.60(3.28) |  |
| Smoke (%) |  |  |  | 0.31 |
| No | 56.01(0.04) | 54.79(1.69) | 57.11(2.09) |  |
| Yes | 43.93(0.04) | 45.21(1.69) | 42.89(2.09) |  |
| Serum albumin (g/L) | 42.97(0.09) | 42.76(0.13) | 43.15(0.10) | 0.003 |
| ACR (mg/g) | 29.17(3.00) | 38.15(5.30) | 21.53(3.85) | 0.04 |
| ACR (mg/g) |  |  |  | 0.001 |
| <30 | 89.87(0.07) | 87.15(0.91) | 92.73(1.06) |  |
| (30-300) | 8.36(0.01) | 10.60(0.85) | 6.51(1.01) |  |
| ≥300 | 1.44(0.00) | 2.25(0.43) | 0.75(0.20) |  |
| Anemia(%) |  |  |  | 0.04 |
| 0 | 93.22(0.07) | 91.65(0.89) | 94.55(0.80) |  |
| 1 | 6.78(0.01) | 8.35(0.89) | 5.45(0.80) |  |
| Hypertension(%) |  |  |  | 0.30 |
| No | 60.77(0.05) | 62.24(2.51) | 59.52(2.46) |  |
| Yes | 39.23(0.04) | 37.76(2.51) | 40.48(2.46) |  |
| DM |  |  |  | 0.05 |
| NO | 83.88(0.06) | 81.20(1.56) | 86.17(2.23) |  |
| Yes | 16.12(0.02) | 18.80(1.56) | 13.83(2.23) |  |
| Hyperlipidemia (%) |  |  |  | < 0.001 |
| No | 30.00(0.01) | 47.43(1.08) | 31.70(1.07) |  |
| Yes | 70.00(0.02) | 52.57(1.08) | 68.30(1.07) |  |
| e-GFR (ml/min/1.73m^2^) | 94.88(0.78) | 96.26(1.05) | 93.70(0.62) | 0.003 |
| e-GFR (ml/min/1.73m^2^) (%) |  |  |  | < 0.001 |
| >60 | 93.77(0.07) | 92.26(1.12) | 95.30(0.77) |  |
| <60 | 6.09(0.01) | 7.74(1.12) | 4.70(0.77) |  |
| RAASi use (%) |  |  |  | 0.001 |
| No | 80.94(0.06) | 77.22(1.75) | 84.26(2.00) |  |
| Yes | 18.96(0.02) | 22.78(1.75) | 15.74(2.00) |  |

DA, docosanoic acid; BMI, Body Mass Index; ACR, albumin-to-creatinine ratio; DM, diabetes mellitus; e-GFR, estimated glomerular filtration rate; RAASi, renin-angiotensin system inhibitors.

**Table 2. Multifactor logistic regression analysis of between DA and the risk of** **CKD in individuals**

| Variables | Unadjusted | | Model 1**^a^** | | Model 2**^b^** | | Model 3**^c^** | |
| --- | --- | --- | --- | --- | --- | --- | --- | --- |
|  | HR (95% CI) | *P*-value | HR (95% CI) | P-value | HR (95% CI) | *P*-value | HR (95% CI) | *P*-value |
| Per-SD increment of DA |  |  |  |  |  |  |  |  |
| Cardiovascular mortality | 0.60(0.36,1.00) | 0.05 | 0.55(0.36,0.85) | 0.01 | 0.55(0.35,0.85) | 0.01 | 0.56(0.35,0.90) | 0.02 |
| All-cause mortality | 0.78(0.62,0.99) | 0.04 | 0.77(0.65,0.91) | 0.002 | 0.76(0.64,0.91) | 0.002 | 0.73(0.59,0.89) | 0.002 |

**Model 1** adjusted for baseline age, sex, ethnicity, BMI; **Model 2** adjusted for covariates in model 1 plus smoke (‘yes’ or ‘no’), alcohol use (‘yes’ or ‘no’); **Model 3** adjusted for covariates in model 2 plus anemia (‘yes’ or ‘no’), hyperlipidemia (‘yes’ or ‘no’), hypertension (‘yes’ or ‘no’), DM (‘yes’ or ‘no’), RAASi use (‘yes’ or ‘no’), ACR, eGFR. DA, docosanoic acid; CKD, chronic kidney disease; HR, Hazard Ratio; CI, confidence interval; BMI, Body Mass Index; DM, diabetes mellitus; RAASi, renin-angiotensin system inhibitors; ACR, Albumin Creatinine Ratio; eGFR, Estimated Glomerular Filtration Rate.

**Figure 1.** Enrollment of flowchart.

**Figure 2.** Association between DA and the risk of CKD in individuals based on restricted cubic spline plot. DA, docosanoic acid; CKD, chronic kidney disease.

**Figure 3.** Associations between DA level and the risk of CKD in individuals. **Model 1** adjusted for baseline age, sex, ethnicity, BMI; **Model 2** adjusted for covariates in model 1 plus smoke (‘yes’ or ‘no’), alcohol use (‘yes’ or ‘no’), poverty, education; **Model 3** adjusted for covariates in model 2 plus anemia (‘yes’ or ‘no’), hyperlipidemia (‘yes’ or ‘no’), hypertension (‘yes’ or ‘no’), DM (‘yes’ or ‘no’), RAASi use (‘yes’ or ‘no’). DA, docosanoic acid; CKD, chronic kidney disease; OR, Odd Ratio; CI, confidence interval; BMI, Body Mass Index; DM, diabetes mellitus; RAASi, renin-angiotensin system inhibitors.

**Figure 4.** Stratified analysis to assess the relationship between DA levels and CKD risk in individuals. Adjust for age, sex, ethnicity, smoke (‘yes’ or ‘no’), alcohol use (‘yes’ or ‘no’), poverty, education, anemia (‘yes’ or ‘no’), hyperlipidemia (‘yes’ or ‘no’), hypertension (‘yes’ or ‘no’), DM (‘yes’ or ‘no’), RAASi use (‘yes’ or ‘no’).

**Figure 5.** Association between DA and the mortality in patients with CKD: (A) Cardiovascular mortality, (B) All-cause mortality. DA, docosanoic acid; CKD, chronic kidney disease.
